# Thalamic deep brain stimulation modulates circadian and infradian cycles of seizure risk in epilepsy

**DOI:** 10.1101/2021.08.25.21262616

**Authors:** Nicholas M. Gregg, Vladimir Sladky, Petr Nejedly, Filip Mivalt, Inyong Kim, Irena Balzekas, Beverly K. Sturges, Chelsea Crowe, Edward E. Patterson, Jamie J. Van Gompel, Brian N. Lundstrom, Kent Leyde, Timothy J. Denison, Benjamin H. Brinkmann, Vaclav Kremen, Gregory A. Worrell

**Author notes:** These authors contributed equally: Vaclav Kremen and Gregory A. Worrell.

## Abstract

Chronic brain recordings suggest that seizure risk is not uniform, but rather varies systematically relative to daily (circadian) and multiday (infradian) cycles. Here, one human and seven dogs with naturally occurring epilepsy had continuous intracranial EEG (median 298 days) using novel implantable sensing and stimulation devices. Two pet dogs and the human subject received concurrent thalamic deep brain stimulation (DBS) over multiple months. All subjects had circadian and infradian cycles in the rate of interictal epileptiform spikes (IES). There was seizure phase locking to circadian and infradian IES cycles in five and seven out of eight subjects, respectively. Thalamic DBS modified circadian (all 3 subjects) and infradian (analysis limited to the human participant) IES cycles. DBS modified seizure clustering and circadian phase locking in the human subject. Multiscale cycles in brain excitability and seizure risk are features of human and canine epilepsy and are modifiable by thalamic DBS.

## Introduction

The unpredictability of seizures is a major cause of disability for people with epilepsy. Despite the apparent randomness of seizure occurrence, in the early 20^th^ century the clinical observations of Sir William Gowers [1] and the study of individuals in the Lingfield Epileptic Colony [2, 3] identified periodic recurrence of seizures for many individuals. More recently, data from ambulatory chronic brain recordings[4-6], patient generated seizure diaries[6], naturally occurring canine epilepsy[7], and mouse models of epilepsy[8, 9] have established that long-timescale cycles of seizure risk are a common feature of epilepsy[10, 11]. These epilepsy cycles operate over multiple timescales with ultradian (briefer than daily), circadian, and infradian (multiday), cycle periods[4, 10-12].

Recent work has further revealed that the periodic circadian and infradian timing of seizures is informed by multiscale fluctuations in the rate of interictal epileptiform spikes (IES), with seizures recurring at preferred phases of IES rate cycles[11]. The complete picture of the multifactorial mechanisms (hormonal, behavioral, brain network) that influence seizure risk remains poorly understood. However, the complex temporal dynamics of seizure risk, including seizure risk cycles, may reflect time-dependent changes in brain excitability, as demonstrated by passive and perturbation-based measures of cortical excitability, network connectivity, and seizure risk[13-21].

The domesticated dog (Canis familiaris) shares an evolutionary history with humans, and dogs are a promising model for the study of human behavior and disease[22-24]. Epilepsy is the most common neurological disorder in dogs[25], and naturally occurring canine epilepsy shares many features with human epilepsy[26]. Additionally, dogs are large enough to accommodate implanted devices designed for humans.

Anterior nucleus of the thalamus deep brain stimulation (DBS) has CE mark and FDA approval for the treatment of drug resistant focal (partial) epilepsy[27]. Prior human and animal studies have demonstrated increased thalamocortical excitability in focal epilepsy[28], and a basic role of the thalamus in the initiation, propagation and continuation of seizures[28, 29]. High frequency DBS has been shown to desynchronize epileptic networks and reduce IES[30]; low frequency cortical stimulation and low frequency stimulation of limbic white matter tracts can reduce brain excitability[31], IES[32], and seizures[33, 34].

Here, we performed long-term (months-to year-long) intracranial EEG monitoring of one human and seven dogs with implantable devices (NeuroVista Seizure Advisory System (NV)[35] and/or investigational Medtronic Summit RC+S™[36]). Three subjects received concurrent thalamic deep brain stimulation (DBS), including high frequency and low frequency DBS settings. We demonstrate that circadian and infradian IES rate and seizure cycles are common in human and canine epilepsy, and provide the first evidence that thalamic DBS produces frequency dependent modulation of cycles in epilepsy.

## Results

### Subjects

Seven dogs (D1-7) and one human (H1) met inclusion criteria. Subjects were monitored with the NV device (n=3 dogs), NV transitioned to RC+S™ (n=2 dogs), or RC+S™ device (n=2 dogs; n=1 human); both devices provide continuous intracranial EEG recordings (iEEG). The RC+S™ device provides both sensing and electrical stimulation. Subjects implanted with the NV device had bilateral subdural strip electrodes spanning the frontoparietal cortices; subjects transitioned from NV to RC+S had custom connectors to interface the RC+S with the NV strip electrodes. Subjects who were implanted exclusively with the RC+S™ device had penetrating depth electrodes targeting bilateral anterior thalamic nuclei (ANT) (Medtronic 3387 or 3389 electrodes) and bilateral hippocampi (Hc) (Medtronic 3391 electrodes). The median recording duration was 298 days (range: 51 – 395), and median seizure burden was 45 seizures (range: 11 – 338) (Supplementary Table 1). D1-5 were monitored in a research kennel, and D6, D7, and H1 (all with RC+S™ system) were monitored in their home environments.

### Chronic brain recordings in human and canine epilepsy

Figure 1a provides a schematic of implanted chronic brain recording devices and shows a representative spontaneous seizure. The running hourly average IES rate is shown for representative subjects (Fig. 1b). Circadian fluctuations in the IES rate showing more frequent spiking during sleep/night are more clearly visualized with a magnified view (Fig. 1c.), and seen as the high frequency oscillation in the hourly average IES rate tracing. Infradian fluctuations in IES rate are evident in the two-day moving average of the spike rate. (Fig. 1b and 1c).

**Figure 1.**
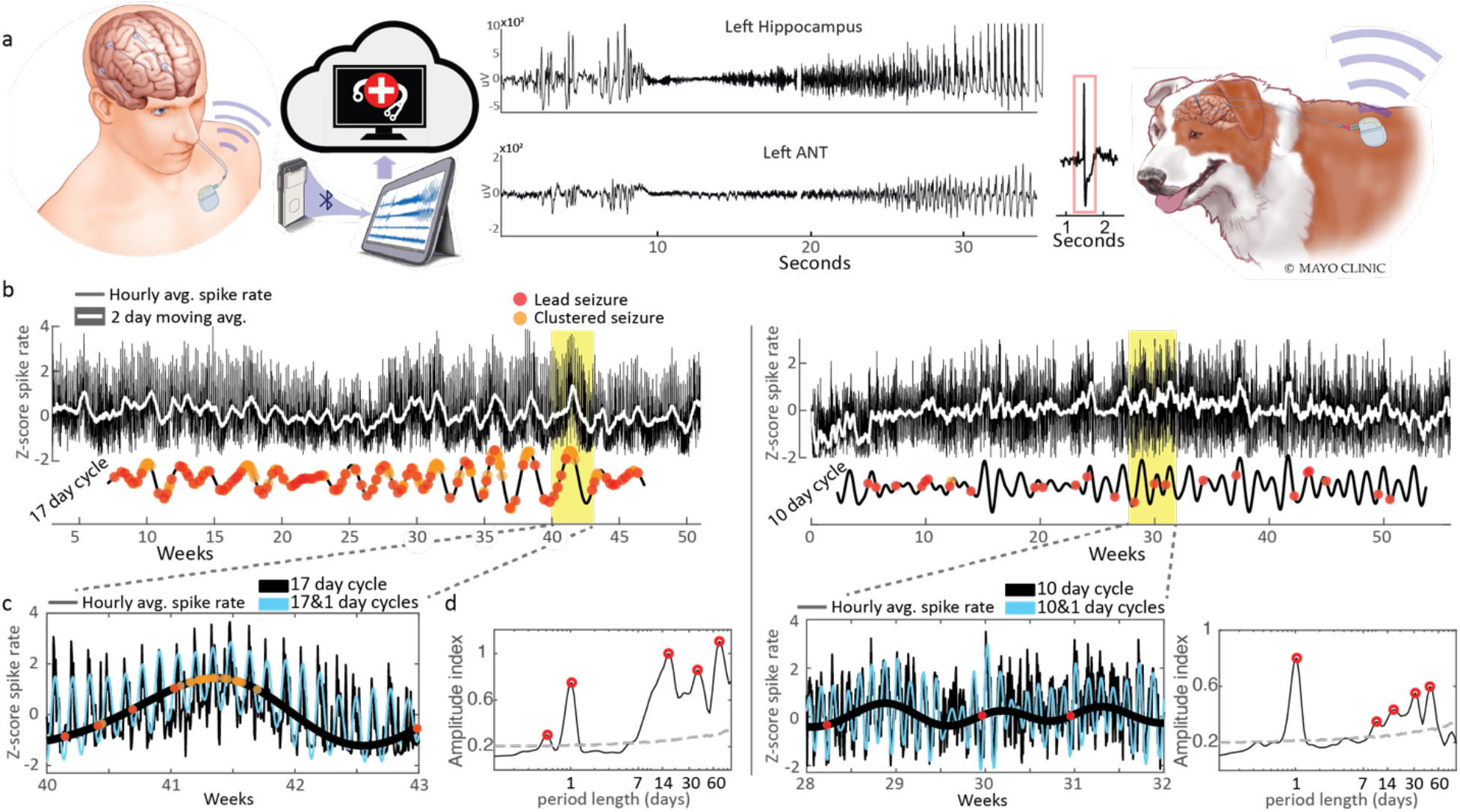
Long-term intracranial EEG of human and canine epilepsy. **a)** Schematic of implanted chronic brain recording devices. **b)** Hourly average IES rate, two-day moving average, and infradian IES rate cycles. **c)** Circadian and infradian IES rate cycles. **d)** Amplitude spectral density. The grey dashed line marks the 95^th^ percentile of normally distributed white noise. Abbreviations: Interictal epileptiform spike (IES)

### Circadian spike rate cycles and seizures

Circadian IES rate cycles were present in all subjects, and are seen as peaks in the wavelet transform of IES rate above the 95% threshold of normally distributed white noise (Fig. 2; Supplementary Fig. 1 and methods for more detail). Figure 2B shows five of 7 dogs and the human subject had 12-hour IES rate ultradian cycles. All subjects had a circadian (∼24 hr) cycle. Interestingly, subject specific infradian cycles were observed in all subjects (further discussion below).

**Figure 2.**
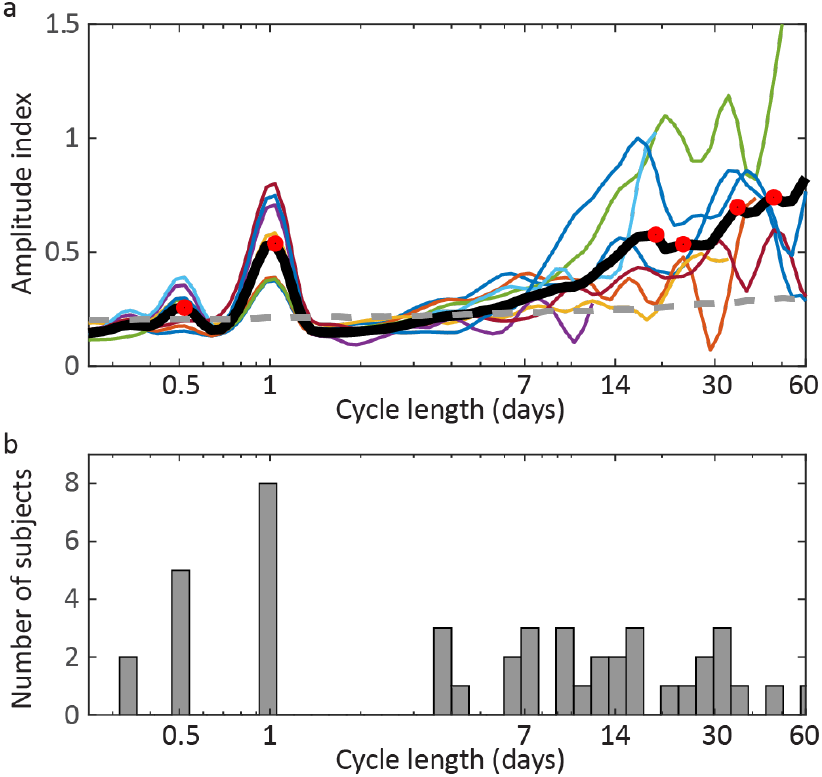
**a)** Hourly average IES rate amplitude spectral density for all subjects and group average. Relative maxima are marked by red circles. The grey dashed line marks the 95th percentile of normally distributed white noise. **b)** Histogram of amplitude spectral density relative maxima above the 95^th^ percentile of normally distributed white noise. The histogram has observed cycles for all subjects (D1-7 and H1). Abbreviations: Interictal epileptiform spike (IES).

Circadian IES rate cycles and seizure timing was evaluated relative to the hour of the day. Figure 3a shows the average normalized daily IES rate, with superimposed seizure onset relative to clock time. Two periods of the averaged cycle are displayed to aid visualization. Twelve-hour IES rate cycles are evident as minor daytime peaks and are most clearly seen in Dogs 6 and 7.

**Figure 3.**
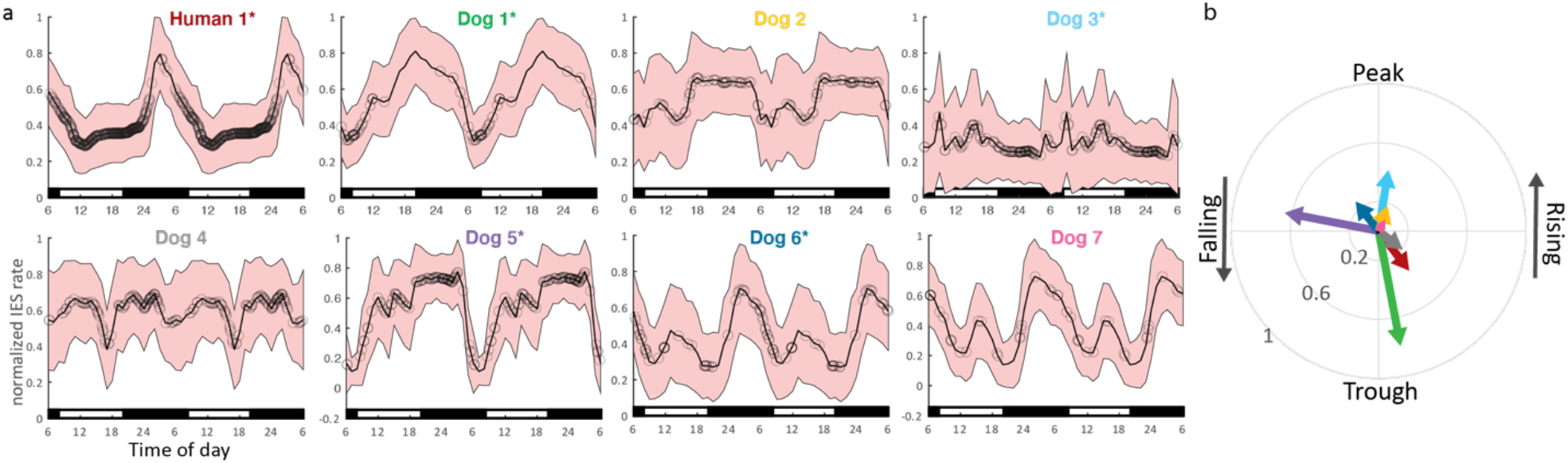
Circadian spike rate and seizure cycles. **a)** Average normalized daily IES rate, with superimposed seizure onset (black circles) relative to clock time. Six hundred and twenty seven total seizures. To aid visualization two periods of the averaged cycle are displayed. Day/night periods are indicated by white/black bars at the bottom of each plot. Twelve-hour IES rate cycles are evident as minor daytime peaks in the canines and are most clearly seen in Dogs 6 and 7. The human subject has a pronounced nocturnal IES peak. **b)** Seizure phase locking to the circadian IES rate cycle. Five subjects had significant seizure phase locking to the circadian spike rate cycle (marked by ‘*’ next to the subject identifier in **a)**). There was no clear group level phase preference of seizure timing relative to the circadian spike rate cycle.

Seizure phase locking to the circadian spike rate cycle is shown in Figure 3b. Five subjects had significant seizure phase locking to the circadian spike rate cycle (marked by ‘*’ next to the subject identifier in Fig. 3a). There was no clear group level phase preference of seizure timing relative to the circadian spike rate cycle.

### Infradian spike rate cycles and seizures

All subjects had significant infradian spike rate cycles (Fig. 2), which were also evident in the 2-day moving average trend of the hourly spike rate (Fig. 1b and 1c). Prior studies have shown inter-individual epilepsy chronotypes, including approximately weekly, 15-day, and 20-day cycle durations[10]. In our cohort, five subjects had significant weekly IES rate cycles, and five subjects had significant 2 – 3 weeklong cycles (Fig. 2). The human subject showed circadian and infradian IES cycles

Significant IES rate cycles identified in Fig. 2 were further evaluated using the filter-Hilbert method, and the filtered infradian cycles reflect slow changes seen in the IES rate 2-day moving average (Fig. 1b). Superimposition of circadian and infradian cycle data provides increasingly representative reproductions of the raw hourly spike rate trend (Fig. 1c).

Seizure phase locking to infradian spike rate cycles was evaluated in subjects with approximately weekly or 2 – 3 weeklong spike rate cycles. Representative circular histograms are shown in Fig. 4b, and the group level resultant vector is represented by a black arrow. Fig. 4c shows the resultant vector of seizure timing relative to spike rate cycles for all subjects with weekly and 2 – 3 weeklong spike rate cycles. The group average vector is shown as a black dashed arrow. Four subjects had significant seizure phase locking to weekly spike rate cycles, and 5 subjects had phase locking to 2 – 3 weeklong spike rate cycles (P<0.05); there was a group level seizure phase preference for the rising phase of infradian spike rate cycles.

**Figure 4.**
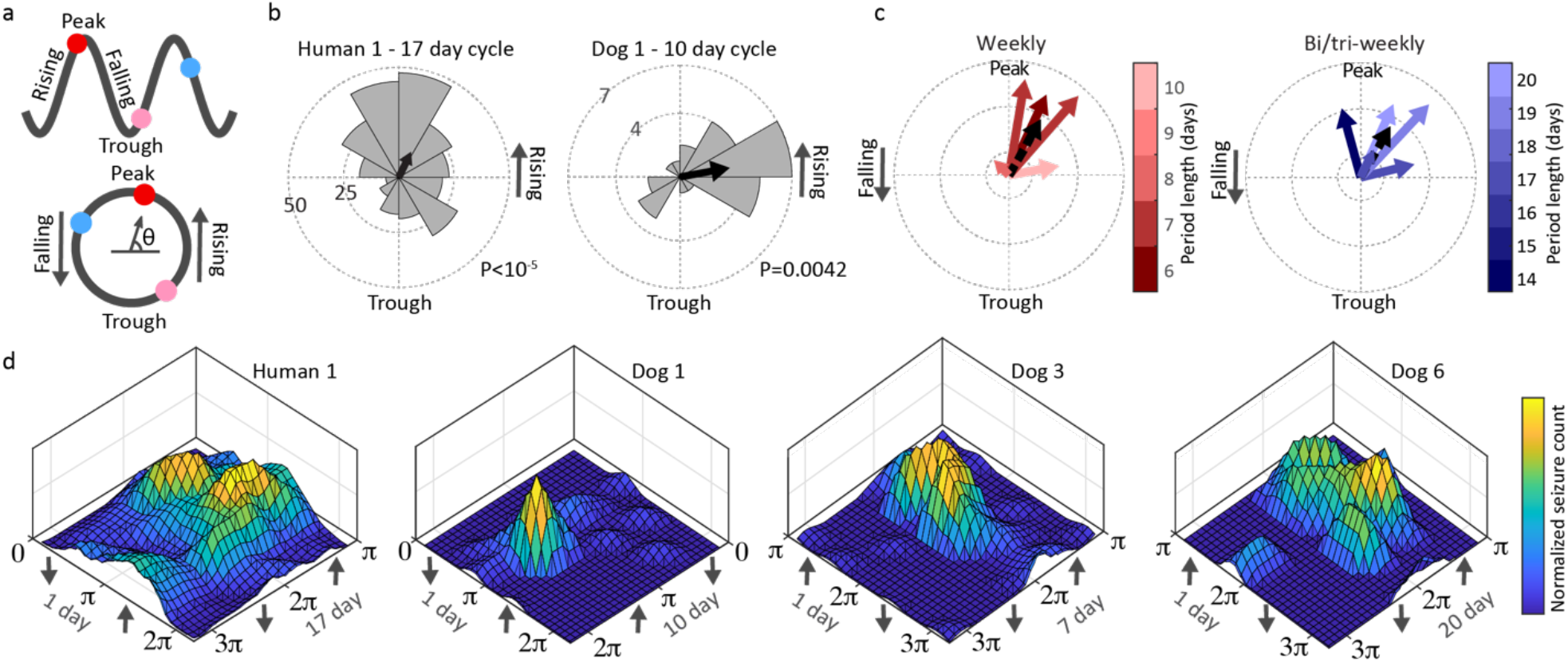
Seizure phase locking to infradian spike rate cycles. **a)** Cycling IES rate depiction showing the increasing and decreasing phases of IES rate. The circle below shows examples of seizures occurring on IES rate late rising phase (red circle), falling phase (blue circle) and early rising phase (pink). **b)** Example of human and dog IES phase diagram showing overall preference of seizures for rising IES rate. **c)** The resultant vector (black dashed arrow group average) of seizure timing relative to IES rate cycles for subjects with weekly and 2 – 3 weeklong IES rate cycles. Four subjects had significant seizure phase locking to weekly IES rate cycles, and 5 subjects had phase locking to 2 – 3 weeklong IES rate cycles (P<0.05). There was a group level seizure phase preference for the rising phase of infradian IES rate cycles. **d)** Phase-phase plots of seizure phase locking to circadian and infradian spike rate cycles shows the interplay of circadian and infradian cycles. As might be expected, infradian cycles are associated with seizures occurring during existing circadian patterns.

### Multiscale spike rate and seizure cycles in epilepsy

Seizure timing was evaluated relative to multiscale spike rate cycles (circadian and infradian). Phase-phase plots show seizure timing relative to the phase of circadian and infradian spike rate cycles for the four subjects with seizure phase locking to both circadian and infradian cycles (Fig. 4). Phase-phase plots show that seizure risk is maximal with circadian and infradian cycles in their respective high risk phase, with declining risk when moving away from the high risk phase of either cycle. Peak risk using phase-phase analyses was 5.8 fold greater relative to infradian cycle-only analysis, and 5.3 fold greater relative to circadian cycle-only analysis for D1; 3.1 and 5.1 for D3; 2.7 and 4.0 for D6; and 2.0 and 1.6 for H1 (Supplementary Fig. 2).

### Thalamic DBS and rhythms in epilepsy

Three community dwelling subjects (H1, D6, D7) living in their natural environment received ANT DBS during the study, in addition to a baseline recording period without DBS. Figure 5a shows the time-frequency spectrogram (CWT scalogram) of hourly average spike rate spanning more than 50 weeks of Hc recordings from H1. Changes to thalamic stimulation were accompanied by clear changes in the amplitude of circadian and infradian (13 – 21 day periods) spike rate cycles.

**Figure 5.**
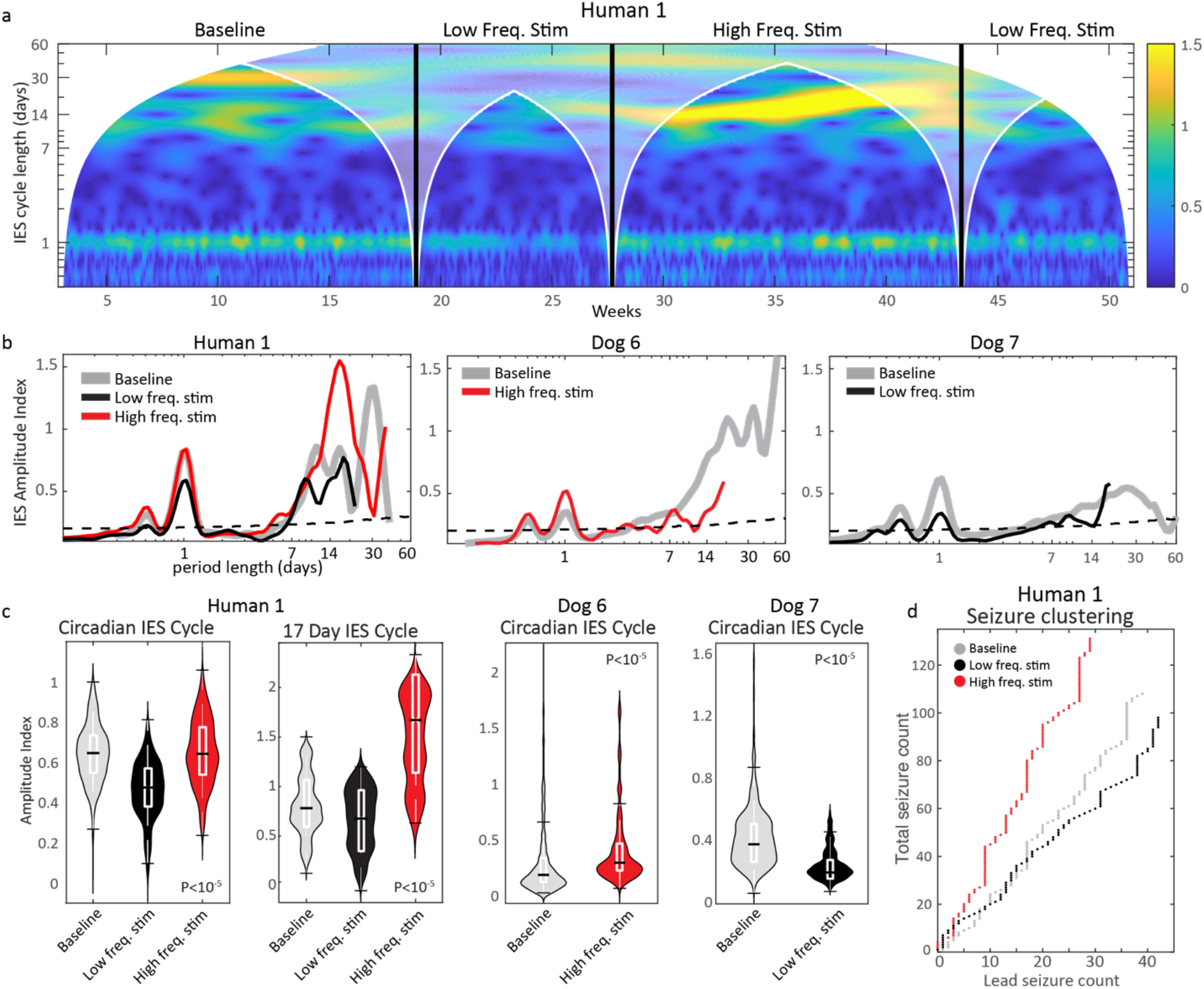
Thalamic DBS modulates IES and seizure cycles in epilepsy. **a)** Spectrogram of hourly average IES rate during DBS-off baseline, low frequency DBS, and high frequency DBS. **b)** Amplitude spectral density of IES rate for pet dogs D6 & D7 and human subject H1. H1 shows frequency specific modulation of the ultradian, circadian and infradian IES rhythms. Similarly, DBS in canines modulated the amplitude of ultradian and circadian rhythms. The impact on infradian canine rhythms was unclear because of inadequate sampling. **c)** Violin plots of circadian and infradian spike rate cycle amplitude in human and pet canines. **d)** Thalamic DBS Also modulates seizure clustering with low frequency ANT DBS reducing overall seizure counts. Abbreviations: Interictal epileptiform spike (IES); Deep brain stimulation (DBS); Anterior nucleus of thalamus.

Following device placement, H1 had 112 days of baseline monitoring with no stimulation, 116 days of low frequency stimulation, and 110 days of high frequency stimulation. The two pet canines living with their owners, Dogs 6 and 7, had 206 and 339 days of baseline recording, and 52 and 0 days, and 0 days and 127 days of high frequency stimulation or low frequency stimulation, respectively (includes recording downtime). A decline in recording adherence during the stimulation-on periods for Dogs 6 and 7 prevented full assessment of infradian cycles with long cycle periods.

The spectral amplitude across stimulation epochs is shown in Fig. 5b. There was significant modulation of circadian (all subjects) and infradian (H1) IES rate cycle amplitude across thalamus stimulation epochs (Fig. 5c; P<10^−5^ for all subjects; statistical significance level was maintained while controlling for the median IES rate for each stimulation state). Interruptions in chronic recordings during thalamic DBS of D6 and D7 limited the duration of infradian cycles that could be assessed, and seizure counts were insufficient for stimulation state subgroup analysis.

Thalamic stimulation had a significant impact on the relative burden of clustered seizures (seizures that occur within 24 hours of a preceding seizure) relative to lead seizures for H1 (Fig. 5d) —there was an increased burden of clustered seizures during high frequency stimulation relative to baseline (Fisher’s exact test P=0.03) and low frequency stimulation (Fisher’s exact test P=0.0009). Furthermore, compared to baseline, there was a trend (nonsignificant) towards increased seizure relative risk with high frequency stimulation (1.18) and reduced seizure relative risk with low frequency stimulation (0.89) (one-way ANOVA P=0.161).

Seizure phase locking to the circadian spike rate cycle was anticorrelated with the amplitude of the infradian spike rate cycle: with low frequency stimulation (and low amplitude infradian spike rate cycle) the circadian seizure phase locking R-value=0.35, P-value 3.7e-6; no stimulation (middle amplitude infradian spike rate cycle) R-value=0.29, P-value 1.2e-4; high frequency stimulation (and high amplitude infradian spike rate cycle) R-value=0.22, P-value=1.3e-3 (phase doubling was used to account for the bimodal circular distribution).

## Discussion

Using long-term brain recordings from novel implantable sensing and stimulation devices in one human and two pet dogs living in natural environments, and five kennel dwelling dogs, we demonstrate that circadian and infradian cycles of seizure risk are features of human and canine epilepsy and can be modified by thalamic DBS. Furthermore, thalamic DBS was shown to modulate epilepsy cycles in a stimulation frequency dependent manner (high vs. low frequency stimulation). To the best of our knowledge this work provides the first evidence that neuromodulation can modulate cycles of seizure risk, suggests that thalamocortical circuits have a role in regulating circadian and infradian rhythms in epilepsy, and expands the evidence that that naturally occurring canine epilepsy can provide valuable insights into the multiscale temporal dynamics of seizures and epilepsy [7].

Circadian IES rate cycles were present in all subjects. Behavioral state dependent changes in IES rate are well established in epilepsy, and sleep/wake related changes likely have a primary role in these cycles. However, beyond behavioral state, there is evidence for circadian cycles in epilepsy dependent on the endogenous circadian pacemaker[37, 38]. Five of eight subjects had seizure phase locking to the circadian IES rate cycle. Recent work based on chronic brain recordings has further found that seizure likelihood varies with day-to-day changes in sleep duration[39]. There was not a consistent inter-subject pattern of the preferred phase of seizure occurrence, consistent with prior human work[10]. We are engaged in ongoing work to sleep stage these chronic recordings to further elucidate the impact of behavioral state on circadian cycles in epilepsy. Five dogs had 12-hour IES rate cycles, which may correspond to daytime naps—naps are a common feature of canine sleep[22, 40]. Interestingly, both community dwelling pet dogs (in contrast kennel dwelling research dogs) had particularly robust ultradian spike rate cycles, which might reflect more stable nap routines.

Infradian IES rate cycles are a well-established feature of human epilepsy[5, 6, 8, 10, 11], and this work provides the first evidence of infradian spike rate cycles in a cohort of dogs with naturally occurring epilepsy (data from D2 was included in a recent review paper on cycles in epilepsy[11]). Infradian chronotypes with approximately weekly and 2 – 3 weeklong cycles have been described in a large cohort (222 subjects) of people with epilepsy undergoing monitoring with a clinical responsive neurostimulation system[10]. All subjects in our study had weekly and/or 2 – 3 weeklong spike rate cycles, with seizure phase locking to these cycles in four and five subjects respectively. Across subjects, seizure occurrences were increased during the rising phase of infradian spike rate cycles, consistent with prior human and rat studies[4, 8]. As others have noted[4], the long period lengths of these cycles preclude characterization during inpatient monitoring stays and may contribute to conflicting descriptions of the relationship between seizures and spike rates.

An important issue to emphasize is that in the absence of a Zeitgeber (from German, ‘time giver’— environmental signals that synchronize or entrain biological rhythms to the external world), endogenous rhythms drift relative to clock time[41]. The gradual desynchronization of a biological rhythm (seizure risk) relative to clock time would limit the usefulness of seizure risk assessments based on a fixed clock time-based cycle. By evaluating seizure occurrences relative to a biomarker of endogenous seizure risk cycles with fine temporal resolution (spike rate), the problem of desynchronization from clock time is avoided. Additionally, by using a composite of neighboring spike rate cycle periods (center cycle period +/- 25%), this method accommodates for fluctuations in the cycle period around a central tendency. These issues highlight the importance of temporally resolved biomarkers of seizure risk, i.e. chronic iEEG. Circadian seizure risk cycles are less disposed to desynchronization from clock time given the presence of powerful daily Zeitgebers, however, desynchronization from clock time could result from shifting behaviors relative to clock time.

Our spike rate and seizure data were generated from a particularly robust dataset of continuous full bandwidth iEEG recordings, using validated spike and seizure classifiers with epileptologist visual review for confirmation. The continuous full bandwidth research recordings support prior human work based on constrained chronic brain recordings from a clinical system[4, 10, 42, 43].

The combination of short (circadian) and long (infradian) timescale cycles can improve the characterization of seizure risk (Fig. 4d)[4]. Such multiscale analyses may hone assessments of seizure risk, reduce the unpredictability of epilepsy, and potentially prompt temporally informed therapeutic and behavioral interventions.

Little is known about the impact of electrical brain stimulation on cycles in epilepsy. In contrast to prior chronic brain recording studies (NV sense only system; Neuropace RNS® system with constrained clinician defined-detectors that are commonly adjusted over time limiting comparisons between detector settings), the RC+S™ device is uniquely suited to evaluate the longitudinal impact of neuromodulation on cycles in epilepsy.

In this study H1, D6, and D7 had bilateral anterior nucleus of thalamus DBS and concurrent bilateral Hc iEEG recording. Circadian spike rate cycles were modulated in a DBS frequency dependent manner—low frequency DBS was accompanied by a reduction in the circadian spike rate cycle amplitude, while high frequency stimulation was accompanied by an increase in circadian spike rate cycle amplitude, in human and canine subjects.

The RC+S™ device requires significant patient and caregiver support to maintain continuous iEEG recordings and streaming telemetry, and several extended gaps of non-recording time during DBS for D6 and D7 required that the data be analyzed in separate chunks, limiting the infradian cycle analysis. H1 maintained reliable iEEG recordings throughout the study, allowing full infradian analysis.

Similar to the modulation of circadian spike rate cycles by thalamic DBS, there was DBS frequency dependent modulation of infradian spike rate cycle amplitude (analysis limited to H1). There was a marked increase in the infradian (17-day) spike rate cycle amplitude during high frequency thalamic DBS, and a reduction in amplitude during low frequency stimulation, when compared to baseline. Additionally, seizure clustering increased during high frequency DBS compared to baseline and low frequency DBS. And finally, seizure phase locking to the circadian spike rate cycle was anti-correlated with infradian spike rate cycles amplitude, with increased seizure circadian phase locking during low frequency DBS (and low infradian spike rate cycle amplitude), and reduced phase locking during high frequency DBS (and high infradian spike rate cycle amplitude), relative to baseline. These findings are consistent with a model of seizure risk in which changes in the amplitude of multiscale spike rate cycles reflects seizure network excitability or seizure risk (Fig. 6). As the infradian seizure risk cycle amplitude increases (Fig. 6 lower panels), there is an increase in the proportion of the circadian cycle that falls above the seizure threshold. This model suggests that the increased amplitude of infradian spike rate cycles, resulting in a larger circadian phase angle above the seizure threshold, would increase the risk of seizure clustering and reduce circadian seizure phase locking, as seen in H1. These findings suggest that beyond seizure phase locking to infradian and circadian spike rate cycles, changes in spike rate cycle *amplitude* may influence seizure risk and be a biomarker to guide epilepsy management.

**Figure 6.**
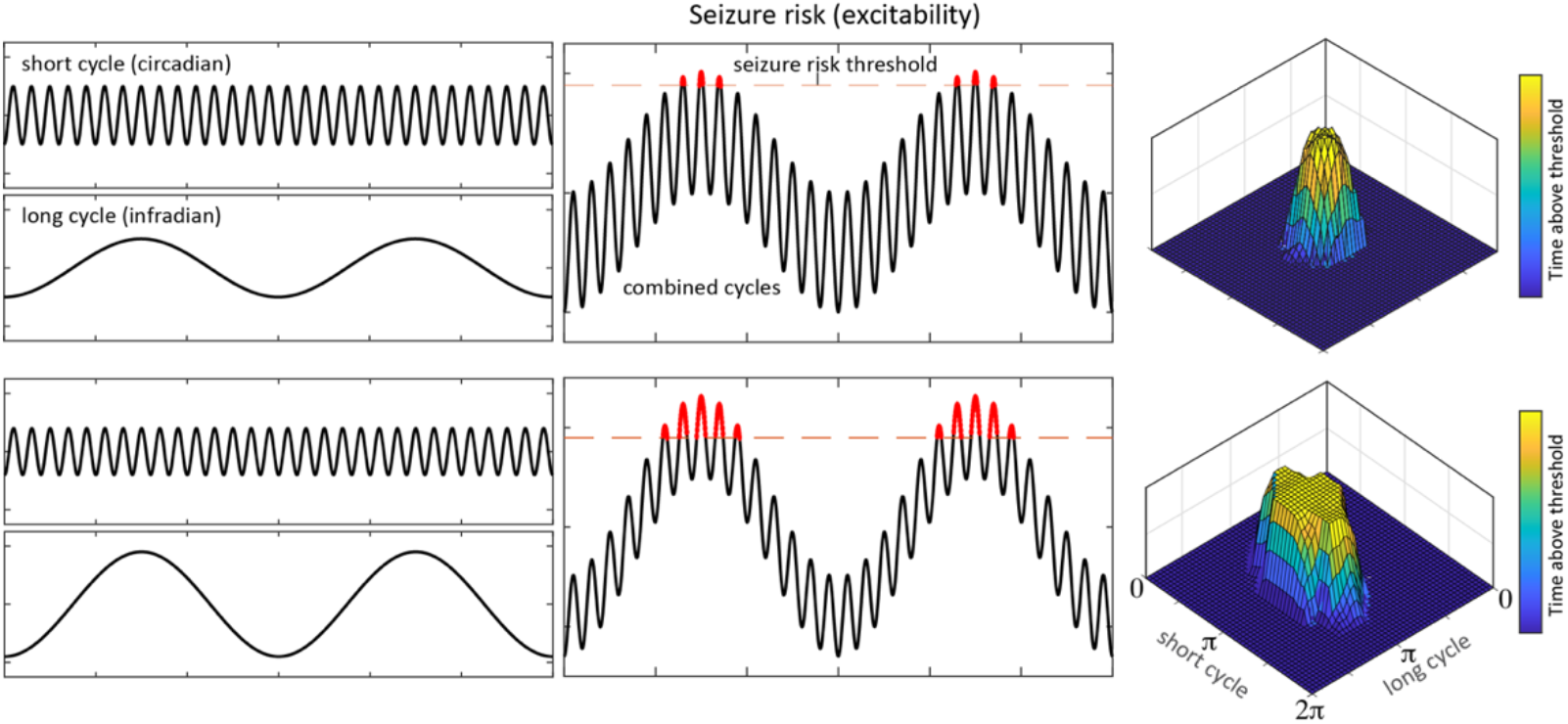
Multiscale spike rate cycle model of the impact of cycle amplitude on seizure phase locking and seizure clustering. The upper panels show short and long cycles of brain excitability independently (left panels), and in combination (center panels). The right panel phase-phase plots mark the phase angle above a theoretical high seizure risk threshold for both the short and long cycles. As the amplitude of the long cycle increases (lower panel), a greater proportion of the short cycle falls above the threshold line, which would result in reduced seizure phase locking to the short cycle. During the high risk phase of the infradian cycle there is an increased risk of seizure clustering as a greater proportion of each day falls above the high seizure risk threshold.

There is evidence that the thalamus plays an important role in focal epilepsy[27], with evidence of enhanced structural and functional thalamocortical connectivity in individuals with temporal lobe epilepsy[44-46], thalamus involvement in the initiation and continuation of seizures[28, 29], and numerous studies showing alterations of thalamocortical circuits in individuals with epilepsy[26, 47, 48]. Here, we provide the first evidence that thalamic DBS can modulates circadian and infradian cycles in epilepsy. This finding raises interesting questions about the role of thalamocortical circuits in the regulation of long timescale dynamics of epilepsy.

DBS is an established therapy for the management of focal epilepsy[27] and various neurological and psychiatric disorders[49], however, DBS mechanisms of action are unresolved[50, 51]. Human and animal studies of high frequency thalamic DBS have demonstrated reduced pathologic epileptic discharges (interictal spikes and high frequency oscillations)[30], increased seizure threshold to chemotoxin[52], and reduced hippocampal synchronization[30, 53]. These investigations relied on acute stimulation protocols, and little is known about the electrophysiological changes associated with chronic stimulation. The pivotal study for ANT DBS[27] delivered high frequency stimulation on a duty cycle (1 minute on, 5 minutes off) without clear supporting evidence; DBS for movement disorders is typically provided using continuous high frequency stimulation. Less is known about low frequency DBS, however there is empirical evidence to support low frequency cortical and subcortical stimulation for the treatment of epilepsy[30, 33, 34, 54, 55], and evidence that noninvasive low frequency stimulation depresses cortical excitability[31].

In our study, subjects received continuous (not duty cycle) high or low frequency stimulation. Continuous stimulation was used in part due to device programming constraints, and was considered acceptable given the established safety of continuous high frequency stimulation based movement disorders protocols, and the lack of a theoretical framework for duty cycle stimulation. Given the increased power in spike rate cycles in H1 and D6 and propensity for seizure clustering with continuous high frequency thalamic stimulation in H1, continuous high frequency stimulation should be used with some caution, and further work is needed to study the different effects of continuous vs. duty cycle stimulation. We did find desynchronization of hippocampal activity and reduced spike rate with acute high frequency stimulation, however, this effect dissipated over time with chronic continuous high frequency stimulation (Supplementary Fig. 3).

This work is limited by the number of subjects with chronic iEEG and concurrent thalamic DBS. Despite the subject count, these chronic recordings form a large dataset with 627 seizures recorded over 2,146 days. Still, this work will benefit from continued iEEG monitoring and additional trials of thalamic DBS. The implantable research iEEG systems in the study require daily charging and device management to maintain connectivity for data transfer, and efforts are being made to ease the burden on participants and improve recording continuity. Seizure cycles were characterized using non-causal filters, consistent with prior studies[4, 6, 10, 42], and future efforts will use causal methods compatible with prospective cycle characterization. We used objective electrographic iEEG seizures in this study, given the unreliability of patient reported seizure diaries and the challenge of tracking clinical seizures in canines. We did not track the behavioral state of subjects in the study so we cannot comment on the role of sleep in cycles in epilepsy, and our group’s automated iEEG based sleep classification efforts are ongoing.

Our results demonstrate that cycles of seizure risk are common features of canine and human epilepsy, which can be modified by thalamic DBS in a frequency dependent manner. This work motivates further study of thalamocortical circuits in the regulation of brain excitability and seizure risk. Naturally occurring canine epilepsy can provide important insights into the temporal dynamics of seizure risk, and next generation implantable sensing and stimulation devices may improve our understanding of cycles in epilepsy and enable new treatment paradigms that leverage the temporal profile of seizure risk and adaptively adjust stimulation to prevent seizures.

## Methods

### Subjects

Nineteen dogs with naturally occurring epilepsy and one human with epilepsy were implanted and monitored with the RC+S™ or NV device (Supplementary Table 1). The human participant and three dogs resided in their natural home environments; the remaining dogs were cared for in a research kennel. Two dogs underwent recording with both the NV and RC+S™ devices in sequence; the lead positions were kept however the leads themselves were exchanged due to cross-device incompatibility. Inclusion criteria required recording of at least 10 electrographic seizures, and greater than 50 days of iEEG. This study was approved by the Federal Drug Administration (IDE G180224) and Mayo Clinic Institutional Review Board.

### Interictal spike and electrographic seizure classification

Data consisted of continuous full bandwidth (250 – 500 Hz sampling rate) iEEG recorded by the NV or RC+S™ systems. Interictal spikes were identified using a validated automated spike classifier[56]. Electrographic seizures were identified by a validated, high-sensitivity automated seizure classifier[57], followed by board certified epileptologist review(G.W.). Hourly IES rates were calculated over consecutive non-overlapping 1-hour blocks of data. Hourly IES rate data were excluded for hours in which data drops accounted for more than 50% of the block. Contacts with inconsistent recordings, artifacts, or lack of seizure involvement were excluded.

### Frequency domain analyses

Frequency domain analyses of IES rate data were performed using the continuous wavelet transform (CWT) and filter-Hilbert transform method. Both of these methods provide instantaneous phase and amplitude information for time-frequency analyses. The CWT was implemented using analytic Morlet wavelets with 10 voices per octave, L2 normalization, and minimum/maximum scales determined by the energy spread of the wavelet in frequency and time. The filter-Hilbert transform method involved narrowband filtering of the broadband EEG signal prior to finding instantaneous phases and amplitudes from the analytic signal generated by the Hilbert transform. Filtering was implemented using least-squares finite impulse response (FIR) bandpass filters of order-3 with zero-phase shift (non-causal). The bandpass center period was incremented by 0.1 days between 0.3 and 2 days, 0.5 days between 2.5 and 10 days, and by 1 day for periods longer than 10 days. The maximum period duration was defined by the recording duration and temporal spread of FIR filters.

When computing the CWT and Hilbert transform, missing hourly IES rate data were filled with random noise drawn from the normal distribution, with mean and standard deviation equal to the distribution of all recorded data from the hour of the day being filled. For long non-recording segments (> 15 days) neighboring records were analyzed independently. For analyses using instantaneous phase/power data from the CWT and Hilbert transform, data within the cone of influence (COI) of boundary effects were excluded (COI defined as 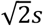 where s is the CWT scale or FIR passband center period—COI shown in Fig. 5A).

### Circadian and infradian spike rate cycles

The CWT was used to assess circadian and infradian cycles in IES rate. Significant cycles were defined as relative maxima in the IES rate CWT amplitude spectral density (ASD) that were above the 95^th^ percentile of white noise. ASD was calculated as the time averaged amplitude of the CWT of the spike rate. To define the 95^th^ percentile threshold of white noise simulations of the CWT ASD (n=1,000) were run on data drawn from the standard normal distribution, and the 95^th^ percentile value was identified for each period duration (Supplementary Fig. 1).

### Seizure phase locking to circadian and infradian spike rate cycles

The CWT ASD was used to identify significant circadian and infradian IES rate cycles. Identified spike IES cycles were further evaluated using the filter-Hilbert method measures of instantaneous phase, and seizure timing was evaluated relative to the phase of these cycles. For each cycle period a composite of FIR filtered IES rate data (spanning period lengths of center frequency +/- 25%) were used to calculate the Hilbert transform analytic signal. This composite cycles data accommodates for drift of endogenous free running rhythms around a central tendency— for example the precise duration of a “monthly” cycle may vary over time around a central trend. Seizure timing was then evaluated with respect to the phase of these circadian and infradian oscillations in IES rate, and circular statistics methods characterized the preferred phase and spread of seizure occurrences relative to IES rate cycles (Fig. 4A), analogous to prior work[4]. Prior work on human epilepsy has demonstrated group level epilepsy chronotypes, including approximately weekly, 15 day, and 20 day cycle durations[10]. Here we emphasized analyses of significant infradian rhythms with weekly and 2 – 3 weeklong cycle periods. For instances with two significant cycles within a period range of interest, the cycle with greater amplitude was selected.

#### Circadian seizure and spike rate cycle

Circadian IES rate and seizure cycles were evaluated using two methods. Method 1 is the same as outlined above, in which a composite of data from neighboring IES rate cycles (centered around the 24-hour period) are used to define the analytic signal for instantaneous phase and power assessments—here, circadian IES rate and seizure data are not evaluated relative to fixed hours of the day, but rather the phase is defined by the circadian oscillation in IES rate itself (may shift from day to day depending on wake/sleep times and other internal and external factors). Circadian cycles are entrained by a powerful exogenous Zeitgeber (24-hour rotation of the earth and the attendant environmental changes). Given this synchronizing Zeitgeber, method 2 evaluated circadian changes in IS rate and seizures relative to the phase of the 24-hour day, and were not defined by the circadian IES rate cycles itself, as in method 1. Method 2 was used for the evaluation of circadian rhythms shown in Figure 3a, otherwise analyses used method 1.

### Circular statistics

Circular statistics provide a statistical approach for the analysis of directional data that are measured on an angular scale—in our case the phase of oscillations in IES rate data. Circular statistics account for the circular character of oscillating or directional data, in which 359° and 1° have nearly the same phase or direction (Fig. 4A). To perform phase analyses of seizures with respect to IES rate cycles, the 0 to 360° phase span (from filter-Hilbert analytic signal) of each period of the IES rate cycle has correspondence to the 360° arc of a circle. The phase of the IES rate cycle at which each seizure occurs can be used to generate circular histograms and visualize the phase preference and spread of seizures relative to oscillations in the IES rate (Fig. 4B). The amplitude of the mean resultant vector, or R-value (also called phase locking value), provides a measure of circular non-uniformity. To calculate the R-value, each seizure is plotted on a circular graph with arbitrary amplitude and phase equal to the phase of the relevant IES rate cycle. Vector summation of vectors drawn from the origin to each event generates the resultant mean vector, and the normalized amplitude (R-value) is a measure of the consistency of event phase. Randomly distributed event will have an R-value of zero, perfectly phase-locked events will have an R-value of 1. For evaluations of circadian seizure timing, the analysis was implanted as outlined above (relative to the circadian oscillation in IES rate), and additionally, seizures were evaluated relative to the time of day.

Statistical significance was assessed by the Rayleigh test[58], which determines the R-value required to reject the null hypothesis that events have a uniform circular distribution. The Rayleigh test assumes that the sample data have von Mises distribution, the circular corollary of Gaussian distribution.

### Phase-Phase plots

For individuals with significant phase locking of seizures to circadian and weekly or 2-3 weeklong IES rate cycles, phase-phase plots were used to further evaluate how combined IES rate cycles inform seizure risk. The circadian and infradian phase data of each seizure was used to generate 3-dimensional linearly interpolated histograms.

### Anterior thalamus stimulation and rhythms in epilepsy

The human participant and two dogs (Dog 6 and Dog 7) underwent chronic ANT therapeutic stimulation. These subjects completed extended baseline “stim-off” recording, followed by extended “stim-on” monitoring. Subjects received bipolar stimulation to bilateral thalami, with the cathodal electrode selected to be positioned within the ANT—stimulation was delivered continuously at low-frequency (<10 Hz) and high frequency (>100 Hz) settings. RC+S™ delivers current clamped stimulation, and stimulation amplitudes ranged between 2 and 6 mA. For the human subject, low amplitude stimulation ≤ 3 mA was considered to be subtherapeutic and was classified as baseline along with stimulation off time periods. The impact of ANT DBS on circadian and infradian IES rate cycles were assessed using frequency domain and circular statistics methods described above. The impact of ANT DBS on seizure rate and seizure clustering was evaluated using statistics described below.

### Statistics

Significant IES rate cycles were defined as relative maxima in the IES rate ASD with amplitudes above the 95th percentile of normally distributed white noise. A one-thousand trial Monte-Carlo simulation was used to define the white noise 95th percentile (Supplementary Fig. 1). For assessments of seizure phase locking to IES rate cycles, circular nonuniformity was assessed by the Rayleigh test[58]. The Rayleigh test determines the amplitude of the resultant vector needed to reject the null hypothesis that events have a uniform circular distribution. One-way analysis of variance (ANOVA) was used to assess for changes in the spectral amplitude of circadian and infradian IES rate cycles relative to thalamic DBS stimulation state. Fisher’s exact test assessed the burden of clustered vs. lead seizures during baseline monitoring and during thalamic DBS (low frequency and high frequency stimulation). All statistics were evaluated at 0.05 significance level. Analyses were performed using MATLAB (version 2020b, Mathworks Inc, Natick, MA, USA).

### Data availability

Data and MATLAB scrips are available from the author by reasonable request.

## Data Availability

Data and MATLAB scrips are available from the author by reasonable request.

## ACKNOWLEDGMENTS

This research was supported by the American Epilepsy Society Research & Training Fellowship for Clinicians (N.M.G.), National Institutes of Health (NIH; U01-NS073557, R01-NS92882, UH2&3-NS95495), and Epilepsy Foundation Epilepsy Innovation Institute My Seizure Gauge. V.K. was partially supported by institutional support from Czech Technical University in Prague. We thank CertiCon a.s. for the use of the Cyber PSG tool. This work benefited from the community expertise and resources made available by the NIH Open Mind Consortium (NIH U24-NS113637; https://openmindconsortium.github.io/).

