## Supplementary Material for "Thalamic deep brain stimulation modulates circadian and infradian cycles of seizure risk in epilepsy"

**Supplementary information**

| Subject | Device | Lead location | Dwelling | Record Duration (days) | Seizure count | Gaps |
|---|---|---|---|---|---|---|
| H1 | RC+S | ANT & Hc | home | 338 | 338 | 14% |
| D1 | NV | frontoparietal neocortex | kennel | 394 | 28 | 21% |
| D2 | NV | frontoparietal neocortex | kennel | 395 | 50 | 14% |
| D3 | NV | frontoparietal neocortex | kennel | 170 | 85 | 1% |
| D4 | NV --> RC+S | frontoparietal neocortex | kennel | 165 | 55 | 17% |
| D5 | NV --> RC+S | frontoparietal neocortex | kennel | 51 | 15 | 7% |
| D6 | RC+S | ANT & Hc | home | 258 | 45 | 37% |
| D7 | RC+S | ANT & Hc | home | 375 | 11 | 52% |

**Supplementary Table 1.** H1 = Human 1. D1 – 7 = Dog 1 – Dog 7. RC+S = Investigational Medtronic Summit RC+S™. NV = NeuroVista Seizure Advisory System. ANT = anterior nucleus of the thalamus. Hc = hippocampus.

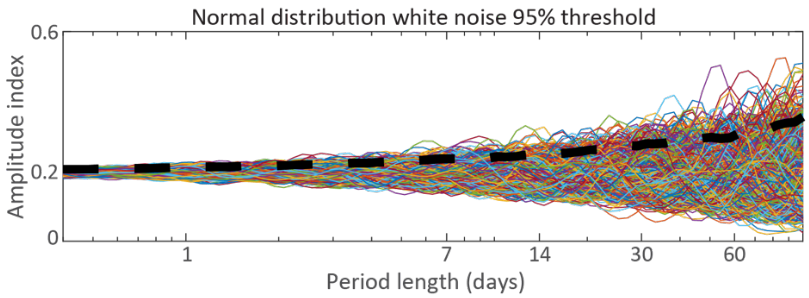


**Supplementary Figure 1.** Amplitude spectral density of n=1,000 simulations of normally distributed white noise, and 95$^{th}$ percentile line.

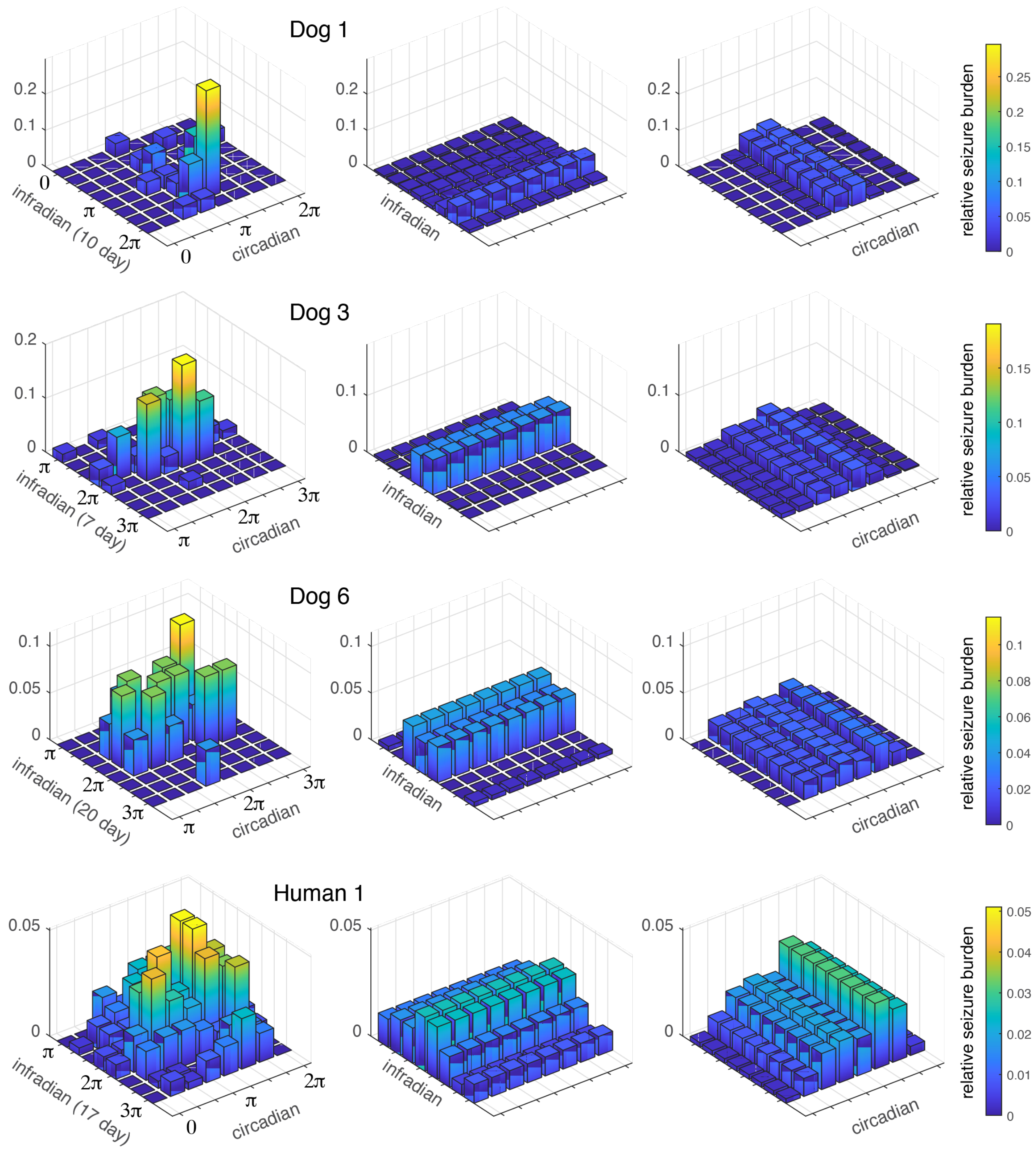


**Supplementary Figure 2.** Seizure risk with phase-phase analysis relative to infradian cycle-only, and circadian cycle-only analysis. Three-dimensional histograms show the burden of seizures in Π/4 x Π/4radian steps, normalized to the total seizure count. For infradian cycle-only and circadian cycle-only plots, seizure counts are constant over the spread of the contrasting (circadian and infradian respectively) cycle.

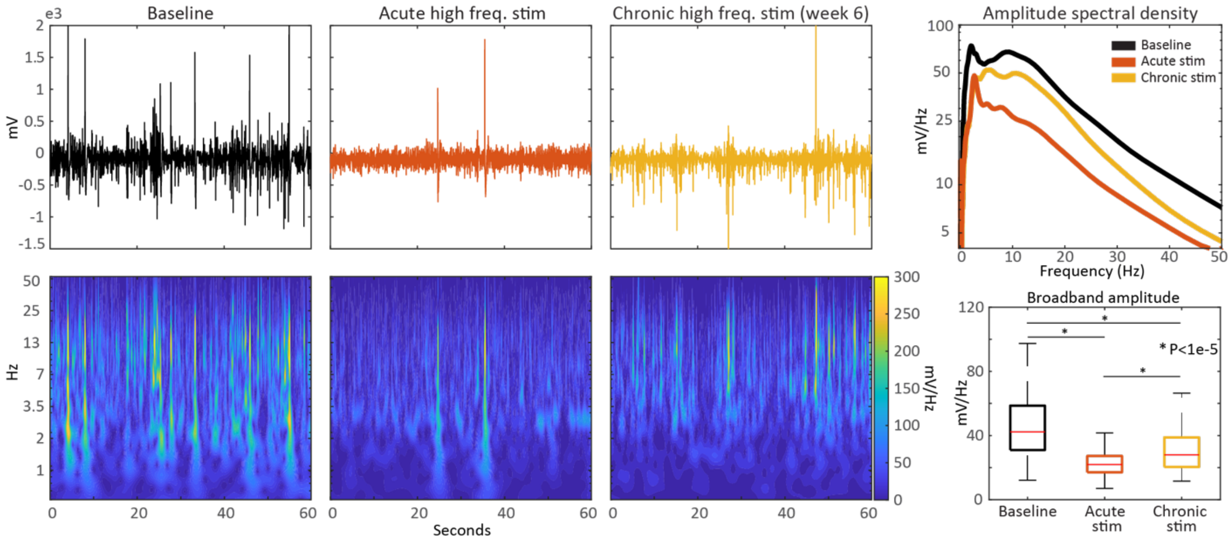


**Supplementary Figure 3.** Representative samples of hippocampal local field potential activity during stimulation off baseline, acute high frequency stimulation, and chronic high frequency stimulation. Broadband amplitude is the 1:50 Hz band mean amplitude.
